# Brain lesion locations associated with secondary seizure generalization

**DOI:** 10.1101/2021.04.19.21255414

**Authors:** Janne Nordberg, Frederic LWVJ. Schaper, Marco Bucci, Lauri Nummenmaa, Juho Joutsa

## Abstract

**Background:** Structural brain lesions are the most common cause of adult-onset epilepsy. The lesion location may contribute to the risk for epileptogenesis, but whether specific lesion locations are associated with a risk for secondary seizure generalization from focal to bilateral tonic-clonic seizures, is unknown.

**Methods:** We identified patients with a diagnosis of adult-onset epilepsy caused by an ischemic stroke or a tumor diagnosed at the Turku University Hospital in 2004-2017. Lesion locations were segmented on patient-specific MR imaging and transformed to a common brain atlas (MNI space). Both region-of-interest analyses (intersection with the cortex, hemisphere, and lobes) and voxel-wise analyses were conducted to identify the lesion locations associated with focal to bilateral tonic-clonic compared to focal seizures.

**Results:** We included 170 patients with lesion-induced epilepsy (94 tumors, 76 strokes). Lesions predominantly localized in the cerebral cortex (OR 2.50, 95% C.I. 1.21-5.15, p = 0.01) and right hemisphere (OR 2.22, 95% C.I. 1.17-4.20, p = 0.01) were independently associated with focal to bilateral tonic-clonic seizures. At the lobar-level, focal to bilateral tonic-clonic seizures were associated with lesions in the right frontal cortex (OR 4.41, 95% C.I. 1.44-13.5, p = 0.009). No single voxels were significantly associated with seizure type. These effects were independent of lesion etiology.

**Conclusions:** Our results demonstrate that lesion location is associated with the risk for secondary generalization of epileptic seizures independent of lesion etiology. These findings may contribute to identifying patients at risk for focal to bilateral tonic-clonic seizures.

## Introduction

Epilepsy is the fourth most common neurological disease, affecting more than 20 million people of all ages worldwide.^1,2^ Brain lesions are the most common cause of adult-onset epilepsy.^3^ Although almost all brain lesions can cause epilepsy, some types of lesions, such as infiltrating tumors, are more prone to cause epilepsy than other types, such as stroke.^3,4^ Clinical risk factors for lesion-induced epilepsy vary depending on the lesion etiology but on a patient level it’s challenging to predict which lesions are likely to cause epilepsy and which are not.^5,6^

Lesion location has been suggested to play a role in the development of lesional epilepsy.^4-6^ In general, lesions in the cerebral cortex are associated with higher risk for epilepsy than lesions in other parts of the brain.^4,5,7,8^ In stroke, lesions in the anterior circulation area are associated with increased risk for late seizures and epilepsy compared to strokes in the posterior circulation.^5,7^ In gliomas, tumor locations in the temporal and frontal lobe have been linked to an increased risk of seizures. However, there is no clear consensus between studies.^4,8-10^ Given that focal seizures can theoretically arise from any brain region, the lack of unifying brain region associated with epilepsy across lesion etiologies is not surprising.

Lesion-induced epilepsy is considered to cause focal epileptic activity, which in some patients spreads to other brain regions outside the focus and may eventually lead to bilateral tonic-clonic seizures (focal to bilateral tonic-clonic, FTBTC).^11^ Secondary seizure generalization in FTBTC seizures involves cortical spreading of epileptic activity to both hemispheres and is hypothesized to include spread to subcortical brain regions including the thalamus, basal ganglia and cerebellum.^12^ However, the underlying mechanisms and pathways leading to FTBTC seizures are still unclear and it is not known if the anatomical lesion location plays a role in the generalization of seizures.^13^ FTBTC seizures are associated with a loss in quality of life^2^, a high risk of injury^14^, and an increased risk for sudden unexpected death in epilepsy (SUDEP).^15^ Identifying lesion locations associated with increased risk for FTBTC seizures may help to identify these patients at an early stage in treatment and benefit patient-specific epilepsy management. Moreover, characterizing lesion locations associated with FTBTC seizures may be valuable for understanding the mechanisms of secondary seizure generalization.^11,16,17^

The aim of this study was to investigate if lesion locations are associated with FTBTC seizures across stroke and astrocytoma, two common lesion etiologies associated with epilepsy. Using both a-priori-region of interest analyses and data-driven voxel-wise analyses, we compared lesion locations of patients FTBTC seizures to patients with focal seizures only.

## Materials and methods

### Patient selection

Patients with lesion-induced epilepsy were identified from clinical records between 2004 and 2017 at Turku University Hospital, Turku, Finland. A systematic database search for patients with age >/=18 years with diagnosis of symptomatic epilepsy (International Classification of Diseases 10, ICD-10 codes: G40.00, G40.01, G40.09, G40.10, G40.11, G40.12, G40.19, G40.20, G40.21, G40.22 and G40.29) and brain MRI obtained up to +/- 3 months from the diagnosis of epilepsy was conducted. A relatively short interval between the epilepsy diagnosis and brain MRI was chosen to minimize the possibility of additional brain lesions or other confounding factors after the occurrence of the lesion. This initial search identified 1887 patients who were selected for a detailed clinical record review to identify cases with new-onset epilepsy, as detailed below.

The case selection is illustrated in **Figure 1**. The clinical diagnoses and seizure types were retrieved from the hospital records. All epilepsy diagnoses were established at the department of neurology at the Turku University Hospital, which is the tertiary care hospital of the Southwest Finland. All relevant clinical records including neuroimaging and clinical neurophysiology examination results and clinical follow up were carefully evaluated by the investigators. Out of 1887 cases evaluated, 748 were confirmed to have focal brain lesions and epilepsy according to the current International League Against Epilepsy (ILAE) definitions.^18^ To ensure we did not include any patients without lesion-induced epilepsy, patients who did have enough information in the hospital records to confirm the diagnosis were excluded. To be able to investigate generalizability of the findings across lesion etiologies, we selected the two most common etiologies in our dataset into this study [astrocytoma (n=201) and ischemic stroke (n=134)]. These cases were selected for a detailed review of the clinical records, including brain imaging and neurophysiological data, to identify patients to the present study.

**Figure 1.**
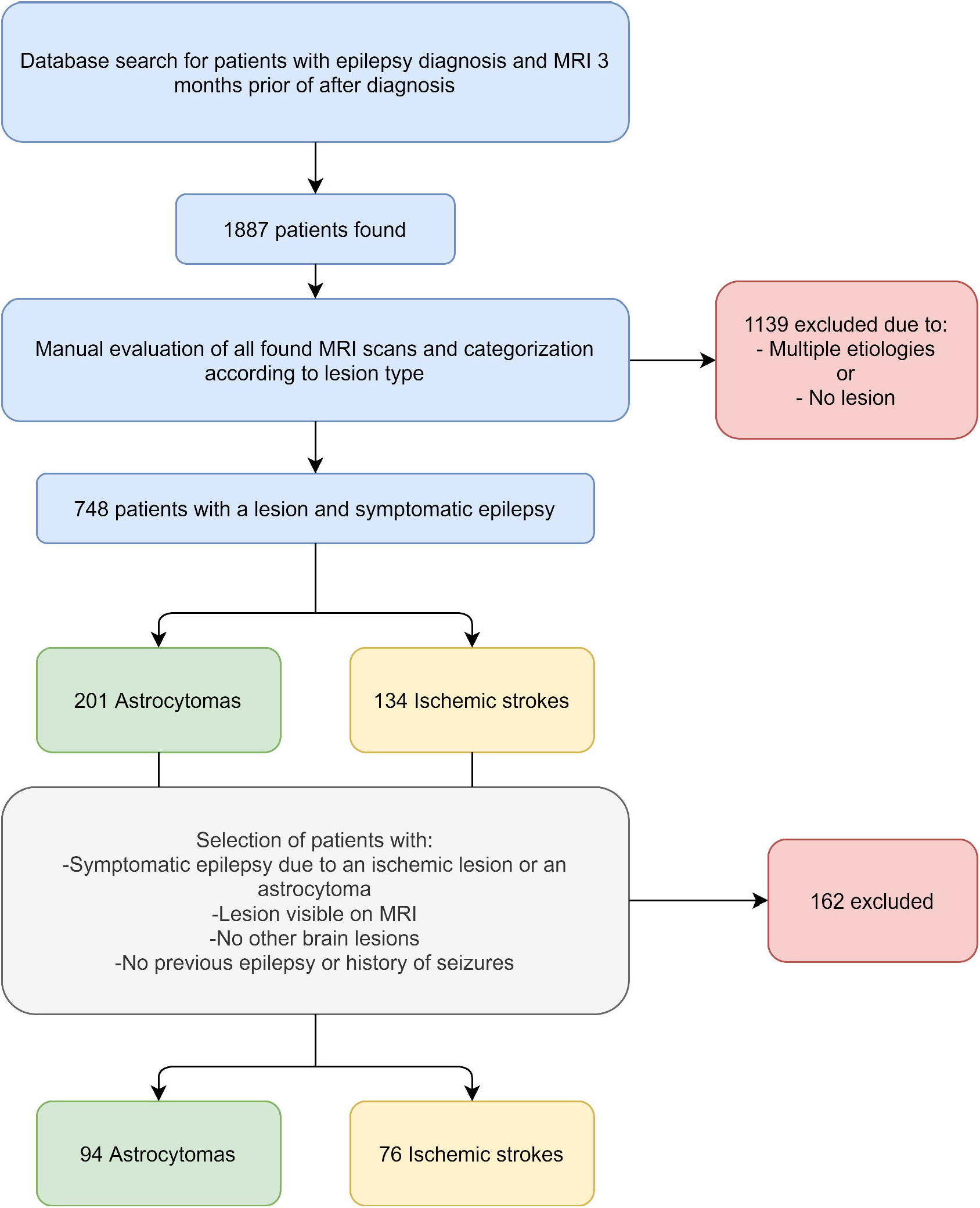
Case selection.

#### Stroke sample

The inclusion criteria were: (i) late seizures consistent with symptomatic epilepsy caused by an ischemic stroke, (ii) one or more ischemic focal stroke lesions in the MRI, (iii) no other brain lesions or structural abnormalities, (iv) no history of epileptic seizures prior to the stroke. Cases with diagnostic uncertainty whether they had focal-onset epilepsy or unclear etiology of epilepsy were excluded. In total, 76 patients fulfilled all criteria and were included in the study (**Figure 1**).

#### Tumor sample

The inclusion criteria were: (i) symptomatic epilepsy caused by an astrocytoma, (ii) one or more tumors with identifiable borders in the MRI, (iii) no other brain lesions or structural abnormalities, (iv) no history of epileptic seizures prior to the astrocytoma diagnosis. Cases with diagnostic uncertainty whether they had focal-onset epilepsy, unclear etiology of epilepsy or evidence of metastases were excluded. In total, 94 patients fulfilled the inclusion criteria (**Figure 1**).

#### Clinical phenotyping

Demographical and clinical data from the selected patients were obtained based on a detailed review of the medical records. Lesion etiology, gender, age at diagnosis, age at first seizure, age at occurrence of the lesion, seizure semiology, EEG findings, number of antiepileptic drugs (AEDs), presence of seizures at best AED therapy (in tumors before any surgery) and status epilepticus episodes were retrieved from the clinical records. In addition, tumor grade (I-IV) was extracted for patients with astrocytomas.

Time of occurrence of the lesion was defined as the time of first symptoms for stroke in the stroke sample and first brain scan showing the lesion in the tumor sample. The epilepsy types were classified to focal only (focal aware or focal impaired awareness), and FTBTC in accordance with the current ILAE classification.^18,19^

#### Lesions

Brain lesions were manually drawn on the high-resolution MR images in the subjects’ native space using FSLeyes software [version 0.31.2, (Jenkinson et al., 2012)]. All slices in the T1 or T2w sequence were examined, and the lesion was approximated on each slice in all three planes. Non-lesioned voxels were assigned value 0 and lesioned voxels value 1, producing a three-dimensional binary lesion mask. The individual MRI was then transformed to the MNI152 standard space using FMRIB’s linear image registration tool (FLIRT) implemented in FSL.^20-22^ Nonlinear registration was not used because structural lesions may affect the gross brain anatomy, leading to bias when using other commonly used nonlinear transformations. Nearest-neighbor interpolation was used to preserve dichotomous voxel classification (lesioned vs not lesioned voxels). The manually drawn lesion mask was registered to MNI152 standard space using the registration matrix obtained from the transformation of the anatomical image to MNI space. The alignment of the transformed images and resulting localization of the lesion in MNI space were manually inspected and edited to ensure accurate anatomical localization of the lesion. Lesion masks were used in both the region-of-interest analyses and voxel-wise analyses.

#### Regions-of-interest

A cerebral cortex mask was defined by combining all cortical regions in the Harvard-Oxford Cortical Structural Atlas with 0.25 threshold.^23^ Cortical involvement was evaluated by quantifying the number of lesion voxels intersecting with the cerebral cortex mask and dividing the number of cortical voxels with the total lesion volume. Lesions with at least half of the voxels located in the cerebral cortex were defined as predominantly cortical lesions. Lesion laterality was evaluated visually using the lesion masks. With bilaterally extending lesions, the hemisphere with the majority of voxels was defined as the predominant side of the lesion. Lobar regions-of-interest (ROIs) were defined in the right and left hemisphere separately to the frontal, parietal, occipital and temporal lobe using the Harvard-Oxford Cortical Structural Atlas by combining all cortical subregions within each lobe. Lesioned voxels in each ROI were calculated and predominantly involved lobe was defined as the lobe with highest number of lesioned voxels.

#### Statistical *analyses*

Differences in demographical and clinical characteristics between samples and epilepsy types were compared using independent samples t-test or Mann-Whitney U-test (continuous variables) and Chi-Square or Fisher Exact test (categorical variables), as appropriate.

Differences in lesion location (laterality and cortical involvement) between cases with focal vs FTBTC seizures in the whole sample were investigated using Chi-Square or Fisher Exact test, as appropriate. In addition, the degree of cortical involvement (percentage of the voxels located in the cerebral cortex) was compared between the groups using independent samples t-test. Finally, a binary logistic regression analysis with seizure type as the dependent variable, and predominant side, cortical involvement and lesion type as independent variables was conducted to study if these factors were independently associated with seizure generalization.

To localize the findings within the hemisphere at lobar-level, ROI analyses were conducted by comparing the number of voxels in each lobe between patients with focal only and FTBTC seizures. The significance of the difference was investigated using Mann-Whitney U tests because the highly skewed distribution of the numbers of voxels in each lobe. Bonferroni correction was used to control for increased type I error due to four lobes included in the analysis. The other hemisphere was used as a negative control to verify the hemispheric specificity of the findings. Independence of the total lesion size was investigated by repeating the analysis with percentages of voxels in an ROI of total lesion voxel, and independence of etiology by repeating the analysis in strokes and tumors separately. Finally, by comparing the proportions of patients with FTBTC seizures in patients with lesions predominantly involving the identified lobe vs. other brain regions using Fisher’s Exact test, and the clinical relevance of the significant findings was quantified by calculating odds ratio for FTBTC seizures for the identified lobe compared to all other lesions using binary logistic regression controlling for lesion type.

### Voxel-based lesion symptom mapping

Voxelwise lesion locations in patients with focal vs FTBTC seizures were compared using voxel-based lesion symptom mapping (VLSM) in both datasets combined. VLSM was conducted using NiiStat software (https://github.com.neurolabusc/NiiStat) according to the published recommendations.^24^ Lesion etiology and lesion size were used as nuisance covariates. The analyses were conducted across the whole brain in voxels affected in at least 5% (n≥9) of the cases and confirmed with thresholds of at least 10% (n≥17) and n≥1. Statistical significance was set at P<0.05 corrected for family-wise error (FWE) using Freedman-Lane permutation with 2000 permutations. The main analysis was repeated in strokes and tumors separately to identify any etiology-dependent effects.

### Ethical statement

The study protocol was approved by Turku University Hospital Clinical Research Services Board, and the study was conducted according to the principles of the Declaration of Helsinki. Due to the retrospective register-based nature of the clinical data collection and large sample size, the need for separate ethical board review and obtaining written consent from the patients was waived.

### Data availability

Voxelwise maps are available upon reasonable request. The original data cannot be made publicly available.

## Results

### Demographics and lesions

The demographical and clinical data of the lesion samples are presented in **Table 1**. There were no differences in the proportion of patients with FTBTC seizures between etiologies (**Table 1**). In patients with focal seizures only, seizures with impaired awareness were more common in strokes compared to tumors (**Table 1**). Focal slowing in EEG was more common in strokes compared to tumors (**Table 1**). Stroke patients were older and had a longer delay between the discovery of the lesion and onset of seizures compared to tumor patients (**Table 1**). Lesion locations were heterogeneously distributed across the brain (Figure 2A) with tumors preferentially localizing to the basal ganglia, insula, temporal lobes and frontal lobes (Figure 2B), and strokes to the regions supplied by the middle cerebral artery (Figure 2C).

**Table 1.** Demographical and clinical data.

|  | Total (n=170) | Tumors (n=94) | Strokes (n=76) | p-value |
| --- | --- | --- | --- | --- |
| **Sex (Male:Female)** | 89:81 | 50:44 | 39:37 | 0.88 |
| **Age at Diagnosis (y)** | 55.9 (17.0) | 47.8 (15.9) | 65.9 (13.1) | **<0.001** |
| **Age at First seizure (y)** | 54.4 (17.0) | 47.1 (16.0) | 63.3 (13.7) | **<0.001** |
| **Age at Lesion discovery (y)** | 53.0 (17.0) | 47.1 (16.1) | 61.0 (14.6) | **<0.001** |
| **Delay from Lesion to seizure (days)** | 462 (1490) | 127 (612) | 978 (2162) | **<0.001** |
| **Seizure type** |  |  |  |  |
| FTBTC seizures | 102 (60.0 %) | 57 (60.6 %) | 45 (59.2 %) | 0.88 |
| Focal seizures only | 68 (40.9 %) | 37 (39.4 %) | 31 (40.8 %) |  |
| ...Focal aware | 41 (24.1 %) | 30 (31.9 %) | 11 (14.5 %) | **<0.001** |
| ...Focal impaired awareness | 27 (15.9 %) | 7 (7.4 %) | 20 (26.3 %) |  |
| **EEG finding** |  |  |  |  |
| Normal | 52 (30.6 %) | 29 (30.9 %) | 23 (30.3 %) | **0.01** |
| Epileptiform | 35 (20.6 %) | 10 (10.6 %) | 25 (32.9 %) |  |
| Focal slowing | 49 (28.8 %) | 29 (30.9 %) | 20 (26.3 %) |  |
| NA | 34 (20.0 %) | 26 (27.7 %) | 8 (10.5 %) |  |
| **Status epilepticus at any time** |  |  |  |  |
| No | 145 (85.3 %) | 84 (89.4 %) | 61 (80.3 %) | 0.10 |
| Yes | 25 (14.7 %) | 10 (10.6 %) | 15 (19.7 %) |  |
| **Seizures on best AED therapy** |  |  |  |  |
| None | 45 (26.5 %) | 30 (31.9%) | 15 (19.7 %) | 0.10 |
| Yes | 122 (71.8 %) | 64 (68.1 %) | 58 (76.3 %) |  |
| NA | 3 (1.8 %) | 0 (0.0 %) | 3 (3.9 %) |  |
| **Number of AEDs in use** |  |  |  |  |
| None | 2 (1.2 %) | 0 (0.0 %) | 2 (2.6 %) | 0.43 |
| 1 | 152 (89.4 %) | 86 (91.5 %) | 66 (86.8 %) |  |
| 2 | 14 (8.2 %) | 7 (7.4 %) | 7 (9.2 %) |  |
| 3 | 2 (1.2 %) | 1 (1.1 %) | 1 (1.3 %) |  |

Mean (SD) or number of subjects (proportion of the respective sample) are shown in the table.
AED = antiepileptic drug. NA = not available. FTBTC = focal to bilateral tonic-clonic.

**Figure 2.**
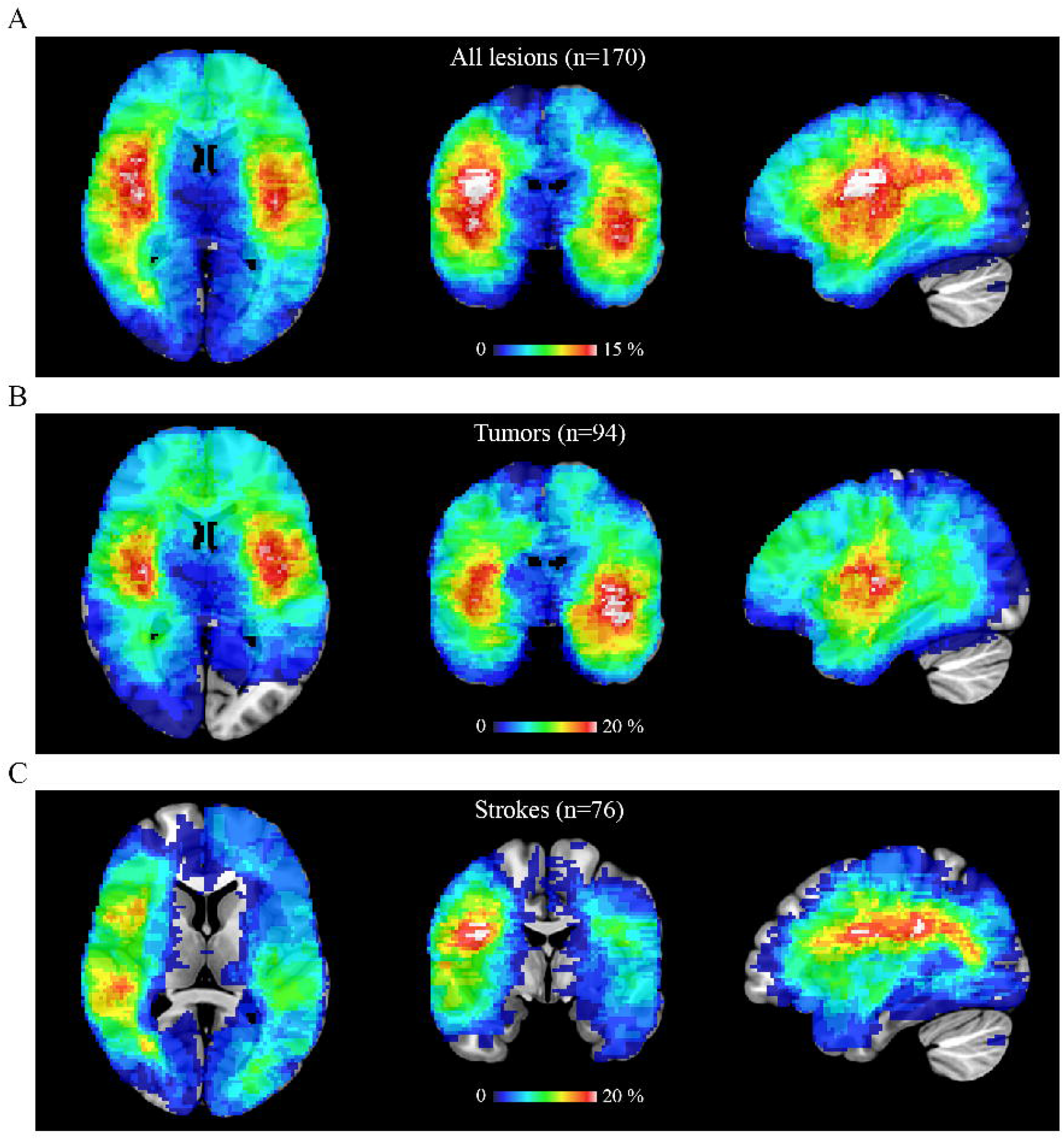
Lesion overlap. Lesion overlap in A) the whole sample (n=170), B) tumor sample (n=94) and C) stroke sample (n=76). The maximum overlap of lesions at any voxel was 18.8%, 28.7% and 23.6% of the sample, respectively.

Patients with FTBTC seizures were slightly younger compared to patients with focal seizures only but there were no other significant differences between the groups in demographic or clinical characteristics (**Table 2**). Demographic data according to the seizure type for the stroke and tumor samples separately are available in the supplementary material (**Table S1** and **S2**, respectively).

**Table 2.** Demographic and clinical data according to the seizure type.

|  | Focal (n=68) | FTBTC (n=102) | p-value |
| --- | --- | --- | --- |
| **Sex (Male:Female)** | 37:31 | 57:45 | 0.49 |
| **Age at Diagnosis (y)** | 58.5 (16.2) | 54.1 (17.8) | 0.10 |
| **Age at First seizure (y)** | 57.0 (16.1) | 52.5 (17.4) | 0.08 |
| **Age at Lesion discovery (y)** | 58.5 (16.1) | 54.1 (17.8) | 0.10 |
| **Delay from Lesion to seizure (days)** | 245 (773) | 595 (1785) | 0.73 |
| **Lesion type** |  |  |  |
| Tumor | 37 (54.4 %) | 57 (55.9 %) | 0.87 |
| Stroke | 31 (45.6 %) | 45 (44.1 %) |  |
| **Lesion characteristics** |  |  |  |
| Size (cm<sup>3</sup>) | 72.1 (73.9) | 79.8 (85.2) | 0.54 |
| Laterality (right-sided) | 28 (41.2 %) | 61 (59.8 %) | **0.02** |
| Cortical | 44 (65.7 %) | 83 (81.4 %) | **0.02** |
| **EEG finding** |  |  |  |
| Normal | 21 (30.9 %) | 31 (30.4 %) | 0.44 |
| Epileptiform | 17 (25.0 %) | 18 (17.6 %) |  |
| Focal slowing | 17 (25.0 %) | 32 (31.4 %) |  |
| NA | 13 (19.1 %) | 21 (20.6 %) |  |
| **Status epilepticus at any time** |  |  |  |
| No | 61 (89.7 %) | 84 (82.4 %) | 0.27 |
| Yes | 7 (10.3 %) | 18 (17.6 %) |  |
| **Seizures on best AED therapy** |  |  |  |
| None | 19 (27.9 %) | 26 (25.5 %) | 0.86 |
| Yes | 48 (70.6 %) | 74 (72.5 %) |  |
| NA | 1 (1.5 %) | 2 (2.0 %) |  |
| **Number of AEDs in use** |  |  |  |
| None | 2 (2.9 %) | 0 (0.0 %) | 0.25 |
| 1 | 60 (88.2 %) | 92 (90.2 %) |  |
| 2 | 6 (8.8 %) | 8 (7.8 %) |  |
| 3 | 0 (0.0 %) | 2 (2.0 %) |  |

Mean (SD) or number of subjects (proportion of the respective sample) are shown in the table.
AED = antiepileptic drug. NA = not available. FTBTC = focal to bilateral tonic-clonic.

### Regions-of-interest analyses

FTBTC seizures were more common in patients with lesions predominantly localized to the cerebral cortex compared to subcortical lesions (65.3% vs 44.2%, p = 0.01, **Figure 3A**), and the right hemisphere compared to the left (68.5% vs 50.6%, p = 0.02, **Figure 3B**). The mean (SD) proportion of voxels of the lesion in the cerebral cortex was 62.0% (19.8) in lesions associated with FTBTC seizures and 54.9% (22.7) in lesions associated with focal seizures only (p = 0.03). The laterality effect remained significant when excluding 21 patients with bilateral lesions from the analysis (68.3% vs 51.4%, p = 0.04). In the logistic regression analysis, both predominantly cortical (OR 2.50, 95% C.I. 1.21-5.15, p = 0.01) and right-sided lesions (OR 2.22, 95% C.I. 1.17-4.20, p = 0.01) were independently associated with FTBTC seizures, while lesion etiology was not (p = 1.0). There were no significant interaction effects between predominantly cortical lesion location, lesion laterality and cortical involvement. Adding age at lesion discovery to the regression model did not change the significance of the effects. There were also no differences in the proportion of predominantly cortical (79.6% vs 69.7%, p = 0.14) or right-sided lesions 51.1% vs 53.9%, p = 0.71) between tumors and strokes.

**Figure 3.**
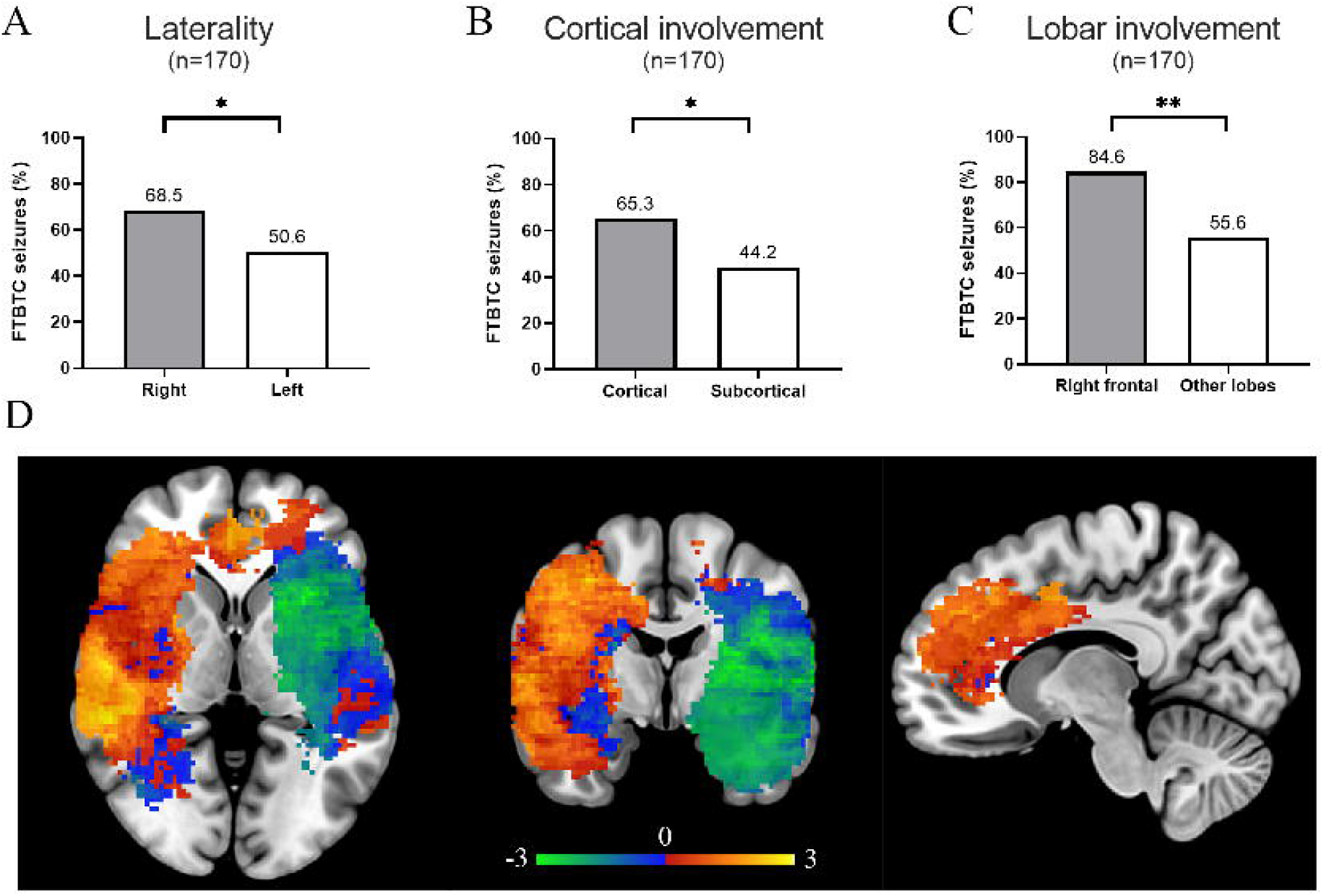
Lesion locations associated with focal to bilateral tonic-clonic (FTBTC) seizures. Proportion of patients with FTBTC seizures according to the predominant side of the lesion (A), predominantly cortical / subcortical lesion location, and involvement of the right frontal lobe (C). Voxel-lesion symptom mapping (VLSM) z map showing brain regions associated with FTBTC seizures (red-yellow scale) and focal seizures (blue-green scale) (D). The sagittal section is from the right hemisphere. There were no voxels significantly associated with the seizure type when corrected for multiple comparisons in the whole search volume.

In the lobar ROI analyses, lesions associated with FTBTC seizures involved the right frontal cortex more than lesions associated with focal seizures only (corrected p = 0.02, **Table S3**). There were no significant differences between the seizure types in other lobes in the right or in any of the lobes in the left hemisphere (**Table S3**). The involvement of the right frontal lobe was also significant when accounting for lesion size (p=0.006), and when analyzing strokes (p=0.048) and tumors (p=0.03) separately]. Lesions predominantly located in the right frontal cortex were associated with FTBTC seizures (OR 4.41, 95% C.I. 1.44-13.5, p = 0.009) compared to lesions located in other regions independent of lesion etiology (p = 0.96) (**Figure 3C**). There also was no significant interaction effect between lesion type and location in the right frontal cortex (p = 0.46). The numbers of predominantly involved ROIs are shown in **Table S4**.

### Voxel-based lesion symptom mapping

The results from the voxelwise VLSM analyses showed clear right-laterization of lesion locations associated with FTBTC seizures (**Figure 3D, Figure S1**). However, the VLSM analysis did not identify any single voxels significantly associated with FTBTC seizures (**Figure 3D**).

## Discussion

The main finding of the present study is that lesion location is associated with secondary generalization of focal seizures (FTBTC seizures). Lesions in the cerebral cortex and right hemisphere, specifically in the right frontal cortex, are associated with an increased risk for FTBTC seizures. These effects were independent of lesion etiology. However, lesions associated with FTBTC seizures did not localize to any single anatomical region within the right frontal cortex, which is in agreement with the current views that seizure generalization is a multifactorial process involving widespread brain regions rather than localized to a single hub in the brain.^11,12^

Previous lesion mapping studies in epilepsy have mainly focused on identification of clinical risk factors for epilepsy and crude anatomical localization of brain regions associated with seizures and epilepsy.^5,7,25^ Across etiologies, lesions in the cerebral cortex are associated with a higher risk for epileptic seizures compared to subcortical lesions, e.g. basal ganglia and cerebellum.^4,5,7,8^ In stroke, lesions in the anterior circulation are associated with increased late seizure risk but result of more detailed localization into which lobes are associated with an increased risk for late seizure have been negative or inconsistent between studies.^26-30^ Studies investigating seizure risk in brain tumors have linked increased seizure risk to varying brain regions, including the medial temporal and frontal lobes.^6,8-10^ However, there doesn’t seem to be consistent evidence on which brain regions are most susceptible for the development of lesion-induced epileptic seizures across etiologies.

The present study focused on identifying lesion locations associated with secondary seizure generalization rather than epileptic seizures *per se*. Our results show that cortical lesions increase the risk for FTBTC seizures, adding to the current knowledge that cortical lesions are more prone to cause any type of seizures.^5,7,25^ Cortical lesions could contribute to the risk of seizure generalization in multiple ways. Greater lesion burden in the cerebral cortex may increase the amount of brain tissue with reduced seizure threshold, causing more widespread or multifocal discharges, or making it easier for epileptic activity to spread from cortical region to another, eventually leading to secondary generalization to FTBTC seizures.^12,31^ Cortical lesions may also lead to a disconnection and loss of cortical control over spontaneous oscillations of subcortical hubs, such as the thalamus, which is known to generate synchronous abnormal neuronal activity in generalized seizures.^11,12,32,33^

Lesion location in the right hemisphere was another independent risk factor for FTBTC seizures in the present study. While previously reported in the literature, the reason for increased risk for secondary generalization of seizures with right hemisphere lesions is not clear. Our finding is in agreement with the results of the large SeLECT study, which reported an increased risk for late seizures associated with right-sided strokes.^30^ The results of our ROI analysis indicated that lesions specifically in the right frontal cortex carry a particularly high risk for secondary generalization. However, similarly to many other studies^6,9,26-30,35^, we did not identify any single common hub within the frontal cortex that, when lesioned, would be significantly associated with epileptic seizures. There are only very few previous studies analyzing the lesion location in a voxel-wise manner and comparing patients with FTBTC to focal seizures. The results from these studies indicate that gliomas involving the corpus callosum^13^ or premotor cortex^8^ could increase risk for FTBTC seizures, but these findings have not been replicated or confirmed in other studies including the present study.

Patients with FTBTC seizures were slightly younger compared to patients with focal seizures only. This is in line with previous observations that younger age is associated with increased risk for late-onset seizures^5^ and could be explained by slow neuroplastic changes, which are known to be more pronounced in younger individuals.^36^ However, age was not independently associated with seizure generalization in the present study and adding age to the regression model did not change the significance of the lesion laterality or cortical location effects.

The strengths of the present study include a focus on seizure generalization and detailed lesion localization by only including patients with high-quality, modern brain MRI with a clearly identifiable focal lesions without mass effects. The analyses were conducted by using several approaches, including a region of interest analyses of crude lesion localization (cortical-subcortical, right-left hemisphere, lobes) and a whole brain voxelwise analysis to identify specific brain regions. Our study was also not restricted to single lesion etiology but included two clinically different lesion types to identify brain regions, which could be important for secondary generalization across etiologies.

There are however some limitations that should be considered when interpreting the results of the present study. First, although our findings indicate that lesion-based localization of FTBTC seizures are independent on lesion etiology, our analyses were limited to only two but clearly different types of lesions (ischemic strokes, astrocytomas). To what extent these findings generalize to across all etiologies, remains to be investigated in subsequent studies. Second, although all of our cases were diagnosed at a university hospital and carefully re-evaluated by the investigators, due to the retrospective nature of the study, it is possible that the clinical characterization of the epilepsy is incorrect in some patients. For example, patients may not report FTBTC seizures because of fearing losing a job or driver’s license. However, incorrect characterization of seizures would lead to increased noise in the data and bias us against the present findings indicating that lesion location is associated with FTBTC seizures. Finally, it is possible that our sample size may not have been sufficient to identify associations between lesions to individual brain regions and seizure generalization. However, if present, such weak associations are likely to have only limited significance in clinical practice.

In conclusion, brain lesions localizing to the right frontal cortex are associated with increased risk for secondary seizure generalization in patients with lesion-induced epilepsy. Our findings can help to better identify patients at risk for FTBTC seizures.

## Acknowledgements

None.

