## Supplementary information for "Brain lesion locations associated with secondary seizure generalization"

**Table S1. Demographic and clinical data according to epilepsy type in strokes**

|  | Focal (n=31) | FTBTC (n=45) | p-value^1^ |
| --- | --- | --- | --- |
| **Sex (Male:Female)** | 16:15 | 23:22 | 0.97 |
| **Age at Diagnosis (y)** | 69.9 (8.8) | 63.1 (14.9) | **0.02** |
| **Age at First seizure (y)** | 67.5 (9.9) | 60.3 (15.1) | **0.02** |
| **Age at Lesion discovery (y)** | 66.6 (11.6) | 57.1 (15.4) | **0.009** |
| **Delay from Lesion to seizure (days)** | 516 (1213) | 1239 (2524) | 0.18 |
| **Status epilepticus at any time** |  |  |  |
| No | 25 (80.6 %) | 36 (80.0 %) | 0.59 |
| Yes | 6 (19.4 %) | 9 (20.0 %) |  |
| **EEG finding** |  |  |  |
| Normal | 10 (32.3 %) | 13 (28.8 %) | 0.96 |
| Epileptiform | 10 (32.3 %) | 15 (33.3 %) |  |
| Focal slowing | 8 (28.6 %) | 12 (26.7 %) |  |
| NA | 3 (9.7 %) | 5 (11.1 %) |  |
| **Seizures on best AED therapy** |  |  |  |
| None | 9 (29.0 %) | 6 (13.3 %) | 0.10 |
| Yes | 21 (67.7 %) | 37 (82.2 %) |  |
| NA | 1 (3.2 %) | 2 (4.4 %) |  |
| **AEDs in use** |  |  |  |
| None | 2 (6.5 %) | 0 (0.0 %) | 0.29 |
| 1 | 27 (87.1 %) | 39 (86.7 %) |  |
| 2 | 2 (6.5 %) | 5 (11.1 %) |  |
| 3 | 0 (0.0 %) | 1 (2.2 %) |  |

**Table S2. Demographic and clinical data according to epilepsy type in tumors**

|  | Focal (n=37) | FTBTC (n=57) | p-value^1^ |
| --- | --- | --- | --- |
| **Sex (Male:Female)** | 21:16 | 29:28 | 0.67 |
| **Age at Diagnosis (y)** | 48.9 (14.7) | 47.0 (16.8) | 0.47 |
| **Age at First seizure (y)** | 48.3 (15.0) | 46.4 (16.8) | 0.52 |
| **Age at Lesion discovery (y)** | 48.4 (14.8) | 46.3 (17.1) | 0.46 |
| **Delay from Lesion to seizure (days)** | 84 (181) | 156 (773) | **0.04** |
| **Status epilepticus at any time** |  |  |  |
| No | 36 (97.3 %) | 48 (84.2 %) | 0.08 |
| Yes | 1 (2.7 %) | 9 (15.8 %) |  |
| **EEG finding** |  |  |  |
| Normal | 11 (29.7 %) | 18 (31.6 %) | 0.12 |
| Epileptiform | 7 (18.9 %) | 3 (5.3 %) |  |
| Focal slowing | 9 (24.3 %) | 20 (35.1 %) |  |
| NA | 10 (27.0 %) | 16 (28.1 %) |  |
| **Seizures on best AED therapy** |  |  |  |
| None | 10 (27.0 %) | 20 (35.1 %) | 0.41 |
| Yes | 27 (73.0 %) | 37 (64.9 %) |  |
| **AEDs in use** |  |  |  |
| None | 0 (0.0 %) | 0 (0.0 %) | 0.65 |
| 1 | 33 (89.2 %) | 53 (93.0 %) |  |
| 2 | 4 (10.8 %) | 3 (5.3 %) |  |
| 3 | 0 (0.0 %) | 1 (1.8 %) |  |
| **Tumor grade** |  |  |  |
| I | 1 (2.7 %) | 2 (3.5 %) | 0.37 |
| II | 16 (43.2) | 22 (38.6 %) |  |
| III | 10 (27.0 %) | 24 (42.1 %) |  |
| IV | 10 (27.0 %) | 9 (15.8 %) |  |

^1^ Mann-Whitney U-test or Fisher Exact Test

**Table S3. Seizure types in lobes**

|  |  | **Focal** |  |  | **FTBTC** |  |  |
| --- | --- | --- | --- | --- | --- | --- | --- |
|  | Mean | Median | IQR | Mean | Median | IQR | p-value^1^ |
| **Right hemisphere lobes** |  |  |  |  |  |  |  |
| Frontal | 680 | 0 | 0-191 | 1502 | 6 | 6-1106 | **0.004** |
| Occipital | 169 | 0 | 0-0 | 176 | 0 | 0-0 | 0.84 |
| Parietal | 569 | 0 | 0-97 | 0 | 921 | 0-675 | 0.09 |
| Temporal | 523 | 0 | 0-29 | 941 | 0 | 0-404 | 0.23 |
| Other | 263 | 0 | 0-61 | 332 | 0 | 0-376 | 0.28 |
| **Left hemisphere lobes** |  |  |  |  |  |  |  |
| Frontal | 1056 | 0 | 0-830 | 1155 | 0 | 0-679 | 0.88 |
| Occipital | 331 | 0 | 0-0 | 173 | 0 | 0-0 | 0.25 |
| Parietal | 711 | 0 | 0-512 | 642 | 0 | 0-105 | 0.18 |
| Temporal | 1077 | 0 | 0-463 | 640 | 0 | 0-3 | 0.081 |
| Other | 432 | 0 | 0-309 | 202 | 0 | 0-53 | **0.029** |

^1^Mann-Whitney U-test

**Table S4. Predominantly involved ROIs**

| **Lobe** | **FTBTC** |  | **Focal** |  |  |
| --- | --- | --- | --- | --- | --- |
|  | Percent | n | Percent | n | p-value^1^ |
| Right frontal | 81.5 | 22 | 18.5 | 5 | 0.025 |
| Right parietal | 66.7 | 18 | 33.3 | 9 | 0.28 |
| Right temporal | 66.7 | 14 | 33.3 | 7 | 0.27 |
| Right occipital | 50.0 | 3 | 50.0 | 3 | 1.0 |
| Right subcortical | 42.9 | 3 | 57.1 | 4 | 1.0 |
| Left frontal | 61.3 | 19 | 38.7 | 12 | 0.44 |
| Left parietal | 47.1 | 8 | 52.9 | 9 | 1.0 |
| Left temporal | 47.8 | 11 | 52.2 | 12 | 1.0 |
| Left occipital | 40.0 | 2 | 60.0 | 3 | 1.0 |
| Left subcortical | 33.3 | 2 | 66.7 | 4 | 1.0 |

^1^Fischer exact test

**Figure S1. Lesion locations associated with focal to bilateral tonic-clonic (FTBTC) seizures** Stroke (A) and tumor (B) lesions associated with FTBTC seizures were predominantly right-lateralized but the voxel-lesion symptom mapping (VLSM) analyses did not reveal any single voxels significantly associated with FTBTC seizures. The sagittal section is from the right hemisphere. The color scales represent z-values.

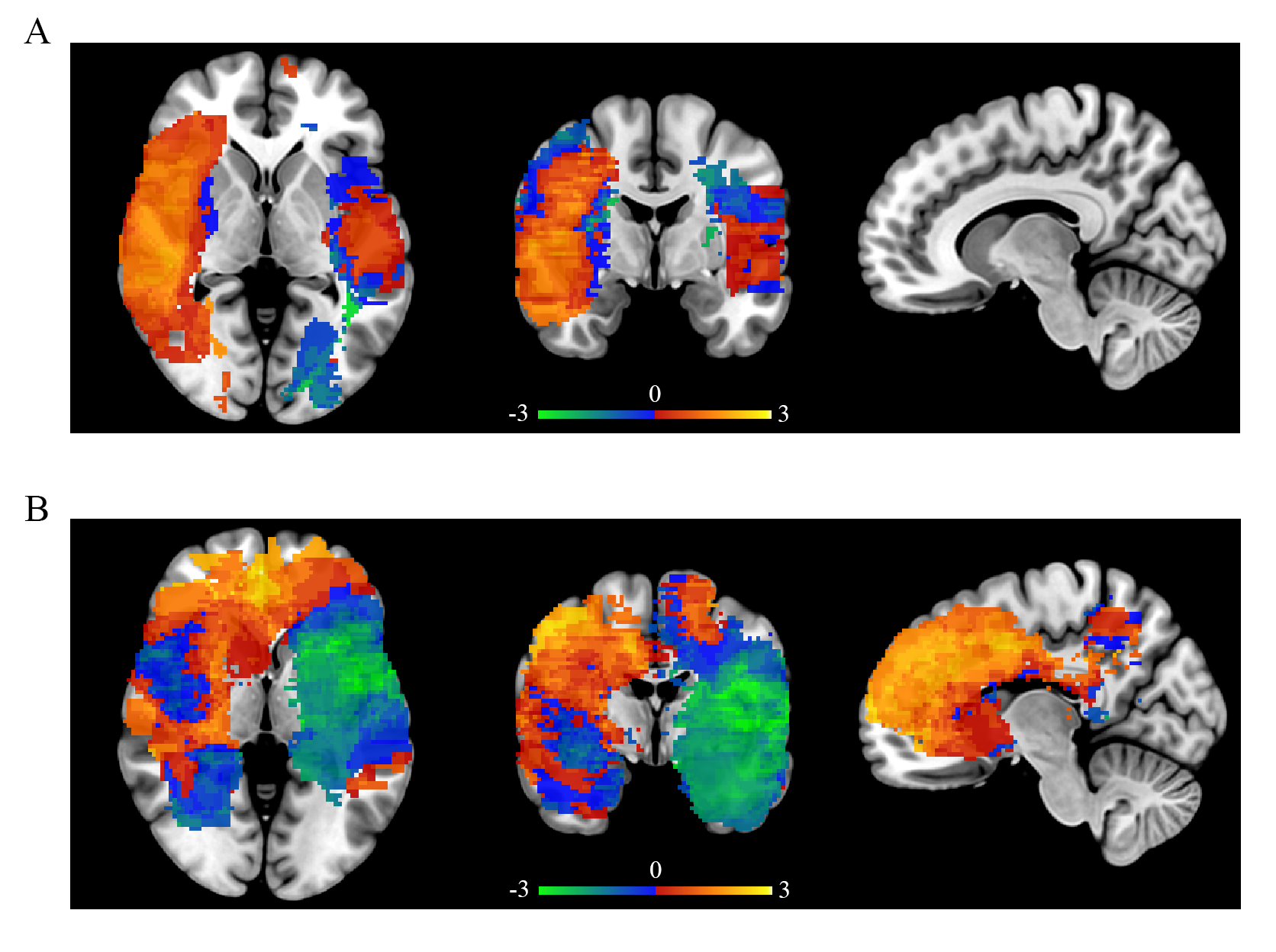
